# BAYESIAN GROUP TESTING WITH DILUTION EFFECTS

**DOI:** 10.1101/2021.01.15.21249894

**Authors:** Curtis Tatsuoka, Weicong Chen, Xiaoyi Lu

## Abstract

A Bayesian framework for group testing under dilution effects has been developed, using lattice-based models. This work has particular relevance given the pressing public health need to enhance testing capacity for COVID-19 and future pandemics, and the need for wide-scale and repeated testing for surveillance under constantly varying conditions. The proposed Bayesian approach allows for dilution effects in group testing and for general test response distributions beyond just binary outcomes. It is shown that even under strong dilution effects, an intuitive group testing selection rule that relies on the model order structure, referred to as the Bayesian halving algorithm, has attractive optimal convergence properties. Analogous look-ahead rules that can reduce the number of stages in classification by selecting several pooled tests at a time are proposed and evaluated as well. Group testing is demonstrated to provide great savings over individual testing in the number of tests needed, even for moderately high prevalence levels. However, there is a trade-off with higher number of testing stages, and increased variability. A web-based calculator is introduced to assist in weighing these factors and to guide decisions on when and how to pool under various conditions. High performance distributed computing methods have also been implemented for considering larger pool sizes, when savings from group testing can be even more dramatic.

## 1. Introduction

The group testing formulation originated by Dorfman (Dorfman (1943)) has found use in a diverse array of applications, including COVID-19 testing. The motivating idea of group testing for a disease like COVID-19 is that, say if biomarker samples from *N, N >* 1 subjects are pooled, and if the prevalence is low, most likely the test result for the pooled sample will be negative, indicating that all *N* subjects in the pool are negative, using only one test. On the other hand, if the result indicates a positive test, and hence there is at least one positive sample present among the pool, then further testing can be conducted to identify the positive subjects. Dorfman suggested that all samples which comprise a positively-tested pool should subsequently be tested individually. This approach has been adopted broadly. Nonetheless, in the presence of testing error, this approach can be problematic, in that classification error thresholds may not be reached. An important potential source of testing error is through dilution, which may occur when pooled samples contain few positives relative to a larger number of negatives. Another important aspect of COVID-19 is the constantly changing prevalence and individual risk levels, such as through seasonality, uneven vaccination rates, and the emergence of new variants. A Bayesian approach is well-suited for acknowledging and incorporating this heterogeneity into the classification process. Described here is a Bayesian framework for systematically addressing these important issues in pooled testing for COVID-19.

Recently, the Food and Drug Administration (FDA) has released guidelines for allowing group testing of patients suspected of having COVID-19. There is thus a renewed flurry of interest in group testing for COVID-19 (Lohse *and others* (2020); Majid *and others* (2020); Hogan *and others* (2020); Shental *and others* (2020); Donoho *and others* (2020)), as it can play a fundamental role in efficient disease surveillance. As there is a large percentage of asymptomatic cases, widespread testing is important for understanding and controlling spread. There also is a need for monitoring large swaths of the population through repeated testing, such as for front line and essential workers, and those at high risk. A potential drawback to such type of surveillance is the reliance on imprecise testing. PCR-based assays based on samples taken with nasal swabs can be highly accurate, although issues with sample collection or mis-priming can lead to errors. These error levels also arise with pooling, and increase with pool size. We refer to this phenomenon as a *dilution effect*. For instance, false negative results from pooling with PCR-based assays that include one positive COVID-19 sample have respectively been estimated to range from 0.07, 0.09, and 0.19 for pools of size 5, 10 and 50 Bateman *and others* (2021). Fast, less expensive, and non-intrusive approaches for testing, such as those based on saliva, can broaden the scope of testing, but may be even less accurate. Emergence of variants also may affect testing error in the future.

A basis for the proposed group testing methods are models known as lattices, in which the classification states that represent the profiles of positivity among test subjects follow a partial ordering. This order structure is used to guide next stage pool selection, and importantly, it provides fundamental insight into the convergence properties of Bayesian group testing with dilution effects. The proposed Bayesian group testing approach explicitly acknowledges testing error and heterogeneity in individual risk levels through prior distribution specification, and can be applied with general response distributions, i.e. either quantitative or categorical test responses. For COVID-19, these outcomes could be viral load level or type of variants detected. Importantly, individual-level classification error can be systematically be reduced to any error threshold, as our proposed approach has attractive optimality properties even in light of dilution effects in which correct classification can efficiently be made for all test subjects. A framework for sequential Bayesian classification on lattices has been described earlier (Tatsuoka and Ferguson (2003)). This current work differs from the previous formulation as we now consider test response distributions that depend on pool size and the number of positives in the pool. In terms of theoretical work involving partially ordered classification models, rules that rely on Kullback-Leibler information of response distributions that are specific to the experiment, such as in cognitive and educational adaptive testing, has been studied (Tatsuoka (2014)). Theoretical properties of group testing problems using partially ordered classification models also have been described (Ferguson and Tatsuoka (2004); Tatsuoka (2005)).

Group testing has a rich statistical literature that has expanded upon Dorfman’s seminal work. An important result in group testing by Ungar (Ungar (1960)) gives a demarcation for when it is optimal to group test when responses are binary and there is no testing error. A Bayesian approach was first described in Sobel and Groll (Sobel and Groll (1966)), where binary responses without testing error are assumed. In addition to screening for disease, group testing has been used in genetics testing (Gastwirth (2000); Sham *and others* (2002)), and to identify promising drug compounds (Xie *and others* (2001); Remlinger *and others* (2006)). The phenomenon of testing error in group testing has been widely recognized (Graff and Roeloffs (1972); Hwang (1976); Litvak *and others* (1994); Thierry-Mieg (2006); Kim *and others* (2007)), including dilution effects with specific functional forms (Hwang (1976); Zenios and Wein (1998); Hung and Swallow (1999)). The possibility that subjects have different probabilities of being positive but with no testing error has been considered (Hwang (1975); Black *and others* (2012)). Beyond binary, responses have been assumed to be normally distributed (Zenios and Wein (1998)), and to reflect viral load levels (Ghosh *and others* (2020)). Group testing has been studied in the context of multiplex and high throughput testing (Bilder *and others* (2019); Donoho *and others* (2020). In (Bilder *and others* (2019)), it is shown that taking into account heterogeneous prior probabilities can improve efficiency. Recently, methods for non-adaptive single-stage COVID-19 group testing have been suggested which are primarily based on the idea of compressed sensing (Shental *and others* (2020); Ghosh *and others* (2020)). These methods have overlapping pools and can potentially identify the positive samples with a single stage of testing. However, they can only work well when prevalence rate is very low (*<* 1.3%), and are not necessarily convergent, as the confounding of positive cases is still possible. There also was no prior information included on samples and no dilution effect. Estimation of proportion such as for disease prevalence is an important application in group testing. Previous work has been based on binary outcomes (Sobel and Groll (1966); Gastwirth and Hammick (1989); Gastwirth and Johnson (1994); Hughes-Oliver and Swallow (1994); Brookmeyer (1999); Tebbs (2003)).

Below, we will first introduce the proposed method and related notions, with an example. We then summarize theoretical properties for optimal designs in Bayesian group testing with general response distributions, and for an intuitive pooled test selection rule. *k*-test look ahead analogues are presented as well, which select *k* tests at a time, and can reduce the back and forth in preparation and testing. Finally, we introduce a web-based tool that implements the proposed algorithms and graphically represents results. Example scenarios are presented, which demonstrate the efficiency gains of group testing, and the trade-offs among the selection rules. As well, a brief note in the Appendix is given on how Bayesian estimation of prevalence can be conducted within the proposed framework through mixtures of posteriors. This is relevant to surveillance of COVID-19. Proofs of theorems and a more thorough technical discussion are also given in the Appendix.

## 2. Methods

### 2.1 Lattice Models for Bayesian Group Testing

We illustrate how lattice-based classification models can be applied in Bayesian group testing with dilution effects.

**Example 1**. Consider all the possible subsets generated by subjects *A* and *B*, as in Figure 1a. Suppose each subset represents a profile of the subjects that are negative for COVID-19. Hence, one of these subsets represents the true state. The collection of these subsets thus form the classification model, and we henceforth refer to them as states: State *AB* denotes that both *A* and *B* are negative, state *A* denotes that only *A* is negative, and hence *B* is positive. Similarly, state *B* denotes that only *B* is negative. State 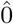 represents that both subjects are positive.

**Fig. 1:**
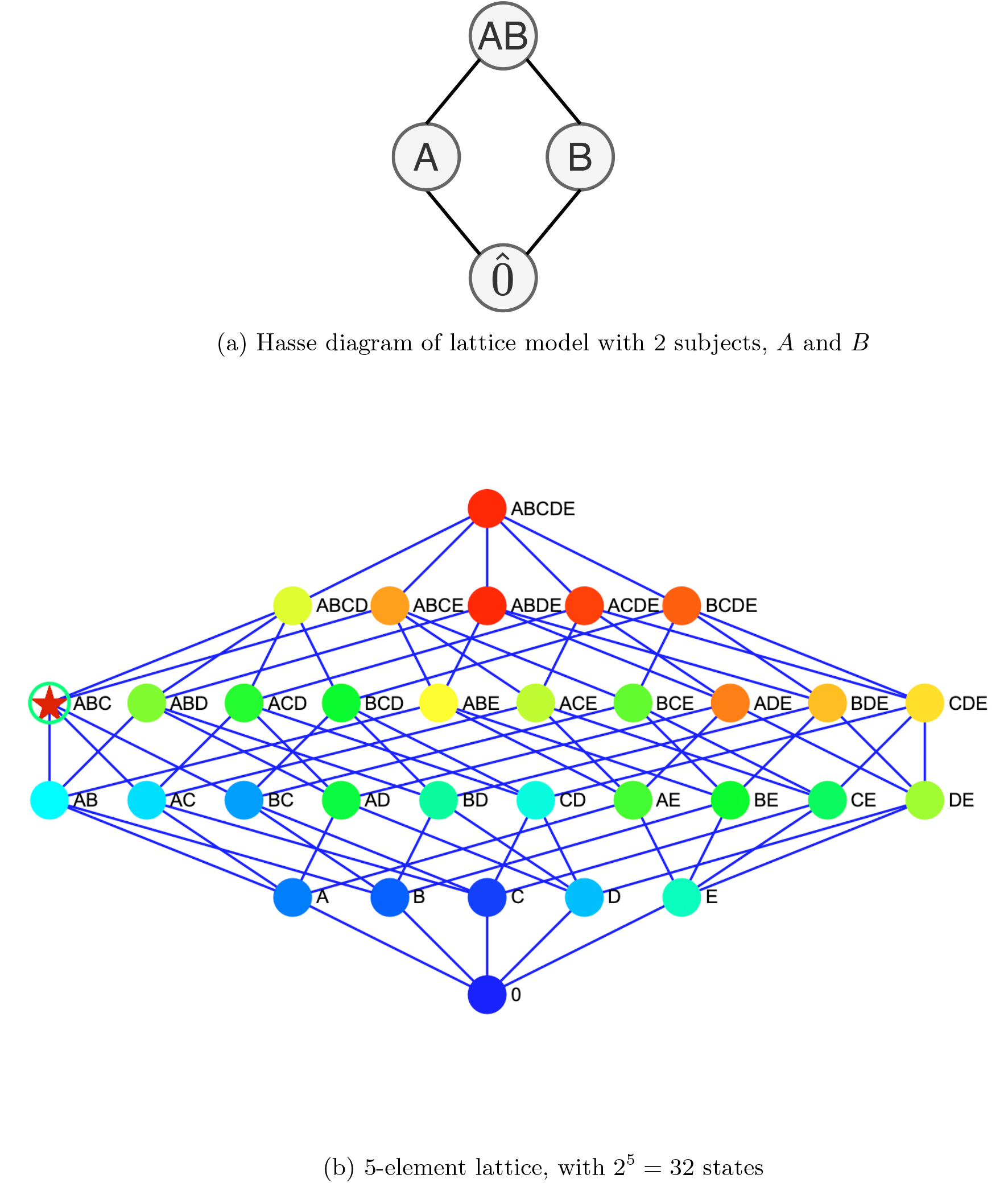
Lattice models

Denote the collection of these states as *S*. In the above example, 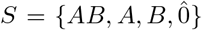. These states (i.e. all the possible subsets of subjects that are negative) actually have a specific order structure in that they can be ordered by inclusion, and form what is known as a powerset lattice. When ordering by inclusion, note for instance that state *AB* is greater than states *A, B*, and 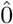 because the set comprised of both *A* and *B* contains *A* alone, *B* alone, and 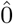, respectively. On the other hand, states *A* and *B* are incomparable, as there is no inclusion relationship between the respective subsets. In this example, there are two subjects, and 4 possible profiles of them being positive or negative.

In general, for *N* subjects, there are 2^*N*^ such possibilities, corresponding to the powerset of the *N* subjects. Note in Figure 1 are lattice models for *N* = 2 and 5 subjects. Further, note that there is essentially a one-to one correspondence between states in lattice model and potential pooled tests. There are 2^*N*^ - 1 possible way to pool tests, since the empty set, represented as 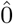, is not a possible test. So, in this example, for state *AB*, there is also a pooled test with samples from both *A* and *B*; for state *B*, there is a test consisting of a sample only from subject *B*, etc. We denote a pooled test associated with a state 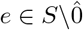 as 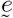.

As the size of classification model and experiment pool grow exponentially in *N*, significant computational challenges arise. We have implemented distributed computing and numerical algorithms that allow models with *N* = 25 (over 33 million classification states and experiments) to feasibly be analyzed when comparing performance across algorithms. We expect this number to further increase with algorithmic and hardware improvements. An overview of computing approaches are given at the end of the Appendix.

We adopt a Bayesian approach. An advantage is the allowance for individual-level prior information about positivity into the classification process, as well as systematic characterization of uncertainty from potential testing error. In a Bayesian framework, the posterior probability values embody the combined empirical and prior evidence as to which state in the classification model is the true one. Larger posterior probability values for the true state are clearly desirable, with a probability value of 1 indicating certainty in correctly identifying the true status of positivity for all the subjects. A *stage* of testing is an iteration of preparing a collection of (pooled) samples for testing and observing the results of the test(s). The posterior distribution after *n* stages of testing on the collection of classification states *S* will be denoted as *π*_*n*_, and for a state *j* ∈ *S*, the posterior probability value that state *j* is the true state is *π*_*n*_(*j*). Corresponding prior distribution values are denoted as *π*_0_ and *π*_0_(*j*).

For instance, prior to testing for COVID-19, each subject could be assigned a prior probability of positivity based on generalized linear regression modeling, with explanatory variables such as age, gender, location of residence, travel history, status of household contacts, community infection rates in relation to specific variants, etc. For a given profile of the *N* subjects in terms of their positivity status, to obtain its prior probability, we take the product of the respective individual prior probabilities for their respective status in the profile. These values form a prior probability distribution across the possible states. After a (pooled) test result is observed, using Bayes rule, prior probabilities are updated to posterior probabilities. We illustrate these computations in the example below.

**Example 1, continued**. Suppose that for a subject *A* the prior probability that he/she is a positive (e.g. has COVID-19) is 0.05, and independently for a higher at-risk subject *B* it is 0.10. The prior probabilities for state membership follows: for state *AB, π*_0_(*AB*) = (0.95)(0.90) = 0.855, for state *A* it is *π*_0_(*A*) = (0.95)(0.10) = 0.095, *π*_0_(*B*) = (0.05)(0.90) = 0.045, and 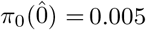. Possible tests include pooling *A* and *B*, and testing each subject individually. Suppose also that a test has two possible outcomes, indicating that either at least one positive is present in the pool, or that only negatives are in the pool. Assume that the specificity, the probability of observing a negative outcome given that the pool does in fact consist only of negative samples, is 0.99. Also, assume that the sensitivity, the probability of observing a positive outcome given that the pool contains at least one positive sample, also is 0.99 when all samples are positive, and reduces slightly to 0.98 when only one of two samples is positive.

Let the first test be comprised of pooled samples from *A* and *B*. Then, if a negative outcome is observed, it can be seen by Bayes rule that *π*_1_(*AB*) = 0.9966. Hence, combined with the prior information, this test result would lead to strong indication that both of the samples are negative. If instead a positive test outcome is seen, then 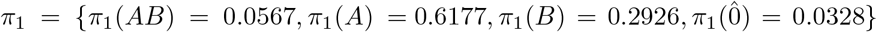. Suppose next that samples from *A* and *B* are tested individually in the next stage as in Dorfman, and the outcome for *A* is negative, and for *B* is positive. If we update the posterior probabilities for the two observed outcomes simultaneously, then, 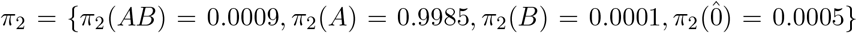. In particular, the posterior probability that *B* is positive is the sum of respective state membership probabilities that indicate that *B* is positive: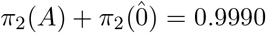. This is clear evidence that indeed *B* is positive. Similarly, note that the probability that *A* is positive is 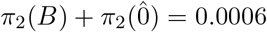, which indicates that *A* is clearly negative.

Making “optimal” pooled test selections becomes more interesting and complex as the number of subjects *N* being considered for pooling increases, the individual risks of positivity vary, and as testing error increases through dilution effects. We consider the following criterion as a basis for optimality in pool selection, from a theoretical perspective. First, let *s* ∈ *S* be the true state (i.e. representing the correct identification of negatives and positives). Typically, 1 − *π*_*n*_(*s*) converges exponentially to 0 at the order *e*^−*αn*^ for some *α*, and *α* is called the *rate of convergence*. The mathematical definition of the rate of convergence is taken to be

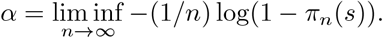

We establish group testing designs and selection rules that attain the optimal rates of convergence (largest possible alpha) with probability one under general dilution effects. Note that rates are in terms of number of tests administered, with one test per stage. We show that they are also characterized in terms of Kullback-Leibler information values, which are discrepancy measures for probability distributions.

### 2.2 Dilution Effects and Optimal Designs

Note that as pool size increases, it is possible that testing outcomes can become less reliable. For instance, for a given number of positive samples, it may be increasingly harder to detect their presence as pool size increases. Another dilution effect can occur in relation to the number of positive samples in a pool. For instance, for a given pool size, it may be increasingly easier to detect the presence of positive subjects as their number increases. Such dilution effects and their effect on the optimality of pooled test selection in terms of attaining the optimal rate of convergence is characterized by Theorem A.1 in the Appendix.

We assume the following conditions that characterize dilution effects, which are formally represented in terms of Kullback-Leibler information values (see Section A.2 in the Appendix). We assume that i) for a given number of positive samples, as pool size increases, pooled tests become less discriminatory in identifying the presence of positive samples; ii) as the difference in number of positive samples between two pools increases for a given pool size, it is relatively easier to discriminate the respective test response distributions; (iii) for a given pool size and a fixed difference in the number of positive samples present, as the total number of positive samples increases, the respective test response distributions become harder to discriminate. Finally, we assume a non-restrictive technical condition, (A4) in the Appendix. Theoretical demarcations when it is preferable to group test depend on which state is true. We illustrate examples of demarcations in the Section A.3 of the Appendix, which show that group testing is attractive asymptotically even under strong dilution effects.

Under these conditions, optimal designs can be considered for three case: 1) all subjects are positive (the bottom state in the lattice), 2) all subjects being tested are negative (the top state in the lattice), and 3) there is a mix of negative and positive subjects (a state in the lattice “inbetween” the top and bottom states). In Theorem A.1, we see that optimal strategies for attaining the fastest possible rates of convergence are: 1) test subjects individually, and 2) to group test samples from all subjects. For the third case, optimal strategies are more complex. The covers and anti-covers of the true state (states directly above and below it) must be considered, as these states pose the most difficulty in discriminating the true state, and hence have posterior probability values that converge to zero the slowest. Under general conditions, the optimal strategy is to group test the negatives and individually test the positives. Optimal rates depend on the statistical discriminatory efficiency of the pooled tests, as measured by Kullback-Leibler information. They also depend on the complexity of the lattice around the true state, as measured by the number of states directly surrounding the true state. Respective tests are selected proportionally so that the rate at which the slowest of the non-true state posterior probability value converges to zero is maximized. In other words, it is desirable that the posterior probabilities of the covers and anti-covers converge to zero at the same rate. More technical details are discussed in Section A.3 of the Appendix. In practice, the true state identity is not known. Next, we propose a testing selection rule that will eventually identify the true state and adopt the corresponding optimal strategy, hence attaining the optimal rate of convergence, no matter the true state.

### 2.3 Bayesian Halving Algorithm and Analogous k-Test Look Ahead Rules

A key to obtaining systematic efficiency gains is to base the sequential selection of pooled tests on the Bayesian information provided by the posterior probability values for the states. In Theorem A.2 of the Appendix, we will establish that under general conditions for dilution effects, an intuitive selection rule for pooling, referred to as the *Bayesian halving algorithm (BHA)*, attains the optimal rate for the true state posterior probability to converge to the value 1. While this is an asymptotic property, given that the rates are exponential, attaining optimal rates is practically attractive for finite testing horizons as well, and insures that any error thresholds will be attained eventually with sufficient testing, regardless of prior specification.

A key concept from order theory is the idea of an up-set for a state in the lattice. This concept is useful for motivating the proposed test selection rules. For a state *j*, denote its up-set as ↑j. This is defined as the subset of states within the lattice that are at least as great as (i.e. contain) the state in question. In Figure 1a, ↑ A*B* = {*AB*}; for *B, ↑* B = {*B, AB*}, for *A,↑* A = {*A, AB*}, and for 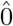 it is all 4 of the states. Practical interpretations of these up-sets are as follows: the up-set of *AB* is all the states for which a pool of *A* and *B* would contain only negative subjects, the up-set of *B* is all the states such that *B* is negative, etc. Conversely, the complement of the up-set of *AB* is defined as 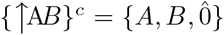, and these are the states for which a pooled test *AB* would contain at least one positive sample. Similarly, 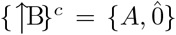 is comprised of the states for which an individual test of a sample from subject *B* would be positive, etc. Hence, the up-set of a state and its complement generate a partition of the classification states according to whether the corresponding pooled test would contain all negatives or not.

Now let us consider rules for pooled test selection based on *π*_*n*_. It is desirable to have a rule that adopts optimal strategies and attains optimal rates of convergence of the true state posterior probability value to 1, no matter which state is true, and under dilution effects, which are likely to arise especially for large pool sizes. The following intuitive pooled test selection rule possesses these properties, as we establish with Theorem A.2 in Section A.4 of the Appendix.

Consider the Bayesian halving algorithm, which selects the pool *e* that minimizes

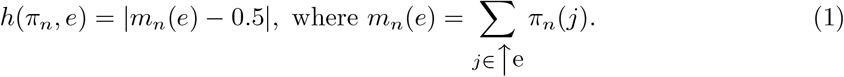

The Bayesian halving algorithm systematically partitions the lattice model, so that regardless of the actual observed outcome, through Bayes rule, one of the two partitions of states will get increased posterior probability values, while the states in the other partition will get decreased values. This systematically encourages posterior mass to eventually accumulate to one state. This property supports that purposeful and systematic pool choices will be made as test results are observed that will lead to correct classification. From Example 1, we compute *h*(*π*_0_, *AB*) = 0.355, *h*(*π*_0_, *A*) = 0.45, *h*(*π*_0_, *B*) = 0.40. Hence, it would be attractive to first select *e* = *AB*, and to group test.

We also extend the halving algorithm to an analogous *k*-test look ahead rule, *k >* 1, so that *k* pools are selected for simultaneous testing, as opposed to the one at a time testing implicit with the proposed halving algorithm. Selecting *k* pooled samples at a time may be more practical, as this can reduce the back and forth involved in preparing pooled samples for testing based on observed results. We define a *stage* of testing to be the simultaneous submission of one or more pooled samples for testing. Given *π*_*n*_ at a stage *n*, the idea behind our proposed *k*-test version of the halving algorithm is to partition the posterior mass on the lattice model into 2^*k*^ partitions according to sets of the following form. We choose *k* pooled samples 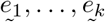 that minimize

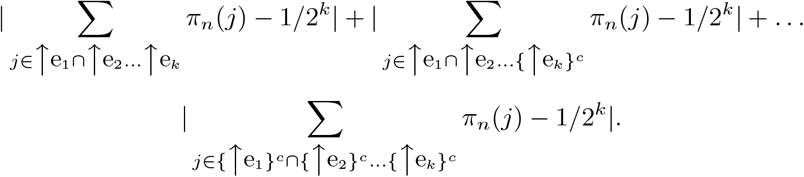

For example, when *k* = 2, this criterion involves selecting the pair 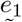 and 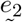 that splits the posterior mass on the lattice model into 2^*k*^ = 4 parts as equally as possible, minimizing

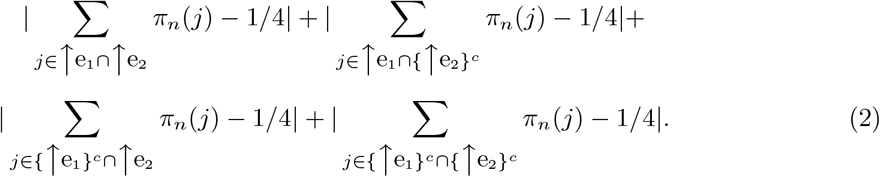

We can approximate the minimization in (2) by sequentially selecting the *k* experiments one at a time. First, given current posterior probability *π*_*n*_, we choose 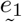 with the halving algorithm, 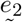 that minimizes (2) given *π*_*n*_ and 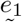, etc. This approximation is computationally much faster.

A halving algorithm that uses prior probabilities is previously described in Black et al.(Black *and others* (2012)), where samples are successively split in half for positive pooled tests while also accounting for heterogeneity in risk. This approach is related to what we propose here, but our approach is fully Bayesian, with posterior probability updating serving as the basis for selection and stopping. Latvik et al.(Litvak *and others* (1994)) present a set of several rules that are related to the halving algorithm. However, the user has to select a rule from this set. These rules differ in the amount of re-testing of pools that is conducted. In contrast, the proposed Bayesian halving algorithm is an automated rule. It also can be used with general test response distributions.

## 3. Results

### 3.1 Simulations

For the Bayesian halving algorithm and its *k*-test look ahead counterparts, *k >* 1, we have created a web-based calculator and code for high performance computing (HPC) distributed computing that generates important performance statistics relating to expected values and standard deviations of number of tests and stages, as well as classification rates, and false positive and false negative rates. The website address is https://bayesgrouptest.case.edu, and the GitHub location for the HPC implementation code is given below. These performance statistics can be used as the basis for comparison and selection of group testing strategies depending on a specific set of conditions. Being able to flexibility assess the trade-offs between different group testing strategies is important for COVID-19 surveillance, as prevalence levels and individual risks, and testing approaches and lab capacities can greatly vary over time and location. Overall performance is derived by averaging state-level expected performance with corresponding state prior probability values for lattice classification. Required inputs are a vector of prior probabilities of positivity for each subject, a matrix with the probabilities indicating presence of a positive that depend on pool size and the number of positives in the pool, and a target posterior probability error threshold for individual classification. Pooled test sequences are also generated. Note that this tool currently only employs binary outcomes for pooled tests (”positive is present”, or not), and henceforth only that case is considered.

**Example 2** We next consider examples with *N* = 12 as an illustration, with varying individual prior probability risk levels: 1) the prior probability value of positivity for each subject is *p*_0_ = 0.02, 2) all such values are 0.20, 3) *m*-mix denotes that *m*-subjects have the prior probability value 0.20, while the rest have the value 0.02. We also assume a dilution effect model modified from Hung and Swallow (Hung and Swallow (1999)), which depends on constants 0 *< γ <* 1 and *h >* 0, the number of positives in a pool 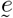, denoted as *r*, and pool size 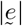. Suppose responses are binary, with specificity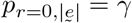, and one minus the sensitivity value

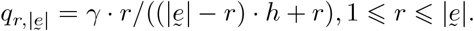

For our examples, we assume *γ* = 0.99, and *h* = 0.005. As an illustration, when pool size is 12, sensitivity ranges from 0.9384 when only one subject is positive, to 0.99 when all of the 12 subjects in the pool are positive. For one positive subject in a pool of size 5, 10, and 50, sensitivity is respectively 0.9759, 0.9529, and 0.7942. These dilution effects are comparable for those found with COVID-19 PCR-based pooling Bateman *and others* (2021). The webtool can generate matrices of dilution effects for varying *γ* and *h*. The dilution effect conditions required in Theorem A.2, which relate to the optimality of the Bayesian halving algorithm, are satisfied with this example. In these settings, we consider performance of individual testing, *k*-test look ahead rules, and the Bayesian halving algorithm (also denoted as *k* = 1).

Finally, stopping of testing is assessed at the individual level. Note that the posterior probability that a subject is negative (positive) is equal to the sum of the posterior probability values of all the states for which the subject is represented as negative (positive). In our example, an individual subject is classified and no longer considered for further testing once this posterior classification error of being negative or positive is at or below a given error threshold. The negative threshold can be stricter to reduce the false negative rate. After subject-level classification, the lattice model is reduced accordingly, mainly to ease computational burden associated with the larger lattice size. Posterior probabilities are renormalized among remaining states. For instance, in Figure 1a, if *A* is removed, then states *AB* and *B* are merged, *A* and 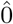 re merged, etc., and posterior probabilities are correspondingly aggregated.

Simulations are based on exhaustive analyses of possible test response sequences, up to a number stages that varies by algorithm. This variation was mainly due to numerical challenges as *k* increases, and computational time limitations. Hence, the reported averages and error levels are approximations. Note for *k* = 1 to 4, we construct simulations up to a maximum of 24, 16, 12 and 11 stages, respectively, and up to 5 stages for individual testing simulations. The overall classification rates by the respective upper stage limit through various prior probability setting of pools (low risks through relatively high risks, non-heterogeneous through heterogeneous) are high, for instance ranging from 99.41% up to 99.99% when the error threshold for stopping is 0.001. For test response sequences that remain unclassified after a maximum stage has been reached, for the computation of performance statistics, it is assumed that the respective test and stage numbers are one more than those last observed. Also, unless stated otherwise, it is assumed that prior probabilities of positivity correctly reflect the risk of the individuals.

Figure 2a shows, as expected, that in terms of average number of tests needed to classify all subjects, group testing with the Bayesian halving algorithm (*k* = 1) has the smallest average number of tests and individual testing the largest, across all the scenarios and selection rules considered here. Indeed, the reduction achieved through group testing can be dramatic. Testing efficiency is particularly striking when the prior probability values of positivity for subjects are small. This gain in testing efficiency reduces as *k* increases among *k*-test look ahead rules. Conversely, as seen in Figure 2b, in terms of the number of stages, individual testing has the smallest average while the *k* = 1 case has the largest. This isn’t surprising given that more tests are submitted per stage as *k* increases. There is thus a trade off between minimizing number of tests versus number of stages, with the intermediate values of *k >* 1 serving as a compromise between the flexibility of single test at-a-time (*k* = 1) group testing versus the relative logistical ease of individual testing. We only consider rules with *k* ⩽ 4 as this still allows for adaptability with respect to test responses, as well as numerical feasibility. Larger values of *k* require greater computational resources in generating and analyzing possible binary testing sequences. The total number of branches grows exponentially, with base 2^*k*^ and exponent equal to the stage number. Figure 2c indicates that the group testing rules can have larger standard deviations for number of tests than for individual testing, and that they increase as prevalence or risk increases, particularly for group testing. Figure 2d indicates that for *k* = 1, the standard deviations for the number of stages are relatively larger, but that as *k* increases, these values become more similar to individual testing. Note that in Figure 2e, the advantage In terms of average false positive and false negative rates, note in Figure 2e that false negative rates appear to decrease, particularly as *k* increases, and as *p*_0_-values increase. We do see relatively larger false negative error for the 0.05 prior for N=12 given threshold 0.005. This is in fact due to the first pool consisting of all 12 subjects, and a negative test that results in immediate stopping for the given threshold. This may be too early. Somewhat elevated false negative rates also arise for *p*_0_ = 0.02 at threshold 0.005, where again stopping is invoked after a first negative test. This holds for 1 ⩽ *k* ⩽ 4. The false negative rate reduces across all prior probability distributions with stricter threshold level of 0.001. Table 1 lists the first stage test selections in different settings of *p*_0_ (again, 1-mix indicates *p*_0_ = 0.02 except for one subject that has value 0.2). Note for instance when *k*=2 and *p*_0_ = 0.02 for all subjects, the first two tests selected for the first stage are to pool all subjects and to individually test subject *A*. For all prior probability scenarios, group testing is initially selected.

**Fig. 2:**
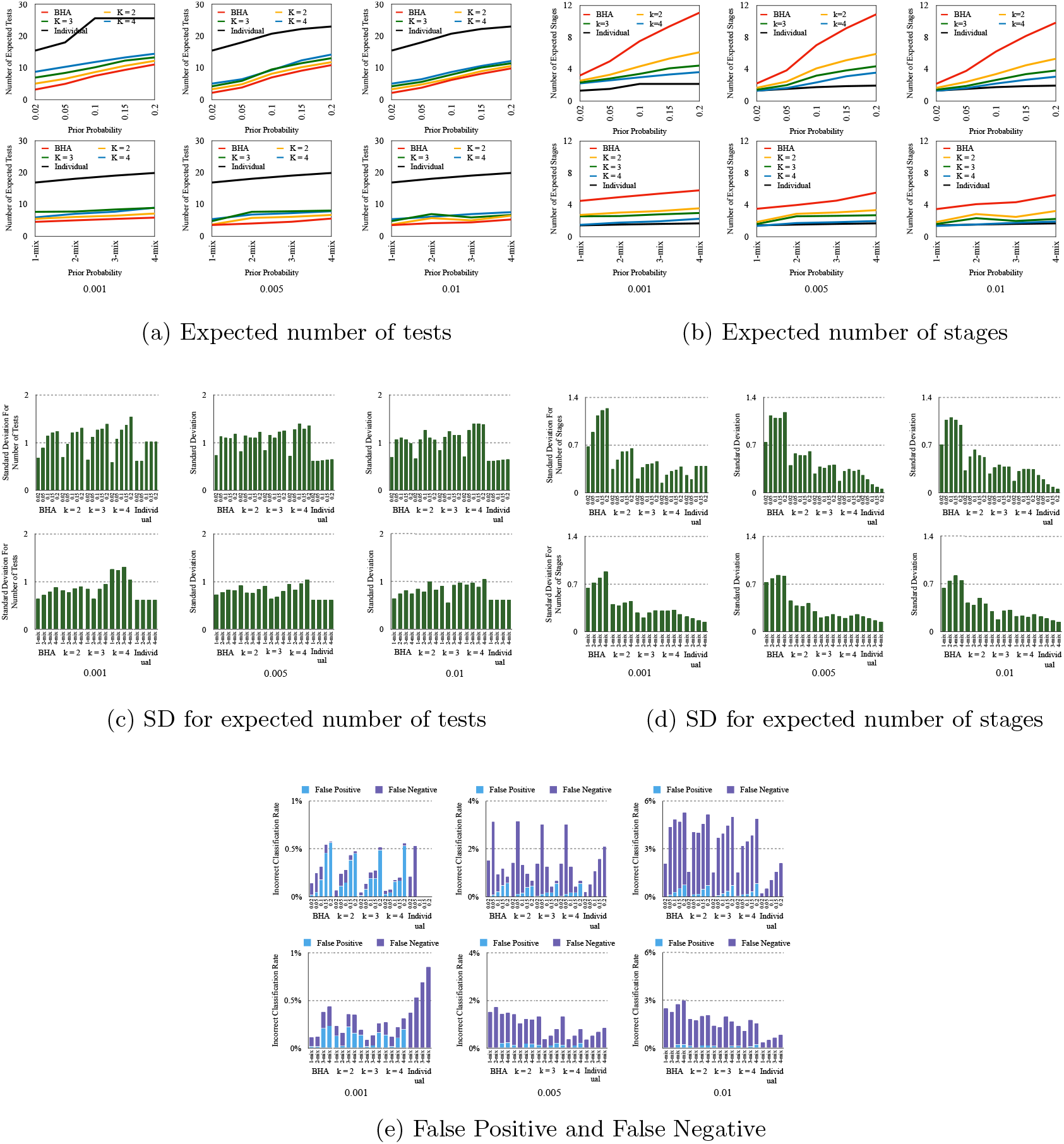
Comparison of expected number of tests and stages, their standard deviations, and false positive/negative rates, using different classification error thresholds for stopping (0.01, 0.005 and 0.001), and under different prior probability scenarios. N = 12.

**Table 1:**
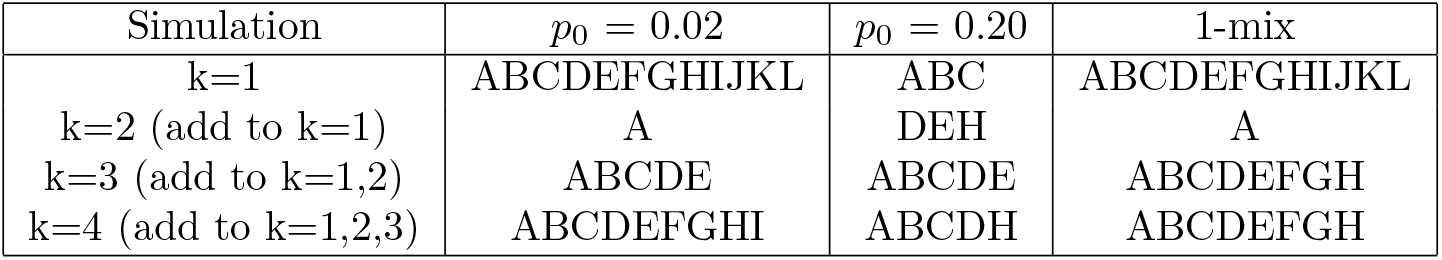
First stage test selections, *N* = 12.

In Figure 3, error threshold is set to 0.01. Note that as N increases, the ratio of expected number of tests for individual testing relative to the proposed group testing rules also increases. For instance, for the case when *p*_0_ = 0.02, as *N* increases from 6 to 16, note that for the Bayesian halving algorithm, this ratio grows from 4.75 to 8.37. The growth rate appears slightly less as *k* increases, with the ratio increasing from 1.47 to 3.63 when *k* = 4. As the prior risk increases, these ratios and the advantage of group testing lessens, as noted in the mixed and *p*_0_ = 0.20 scenarios. For instance, for the Bayesian halving algorithm, the ratios range from 1.77 to 2.36, and for *k* = 4, 0.99 to 1.74. Conversely, the ratio of the expected number of stages with group testing versus individual testing also increases in *N*. For instance, for the case when *p*_0_ = 0.02, as *N* increases from 6 to 16, note that for the Bayesian halving algorithm, this ratio increases from 1.26 to 1.91. Interestingly, the rate of decrease appears slower as *k* increases, with a ratio equal to 1.37 for *N* = 16 when *k* = 2. As prior risk increases, these ratios are larger, and the relative “cost” of group testing increases. Note when *p*_0_ = 0.20, for the Bayesian halving algorithm, the ratios range from 7.72 to 15.93. For *k >* 1, the ratios of expected stages are smaller. For *k* = 4, ratio values range from 4.44 to 6.76. Roughly, this trade-off appears more attractive for lower prior risk, and less attractive as prior risk levels increase.

**Fig. 3:**
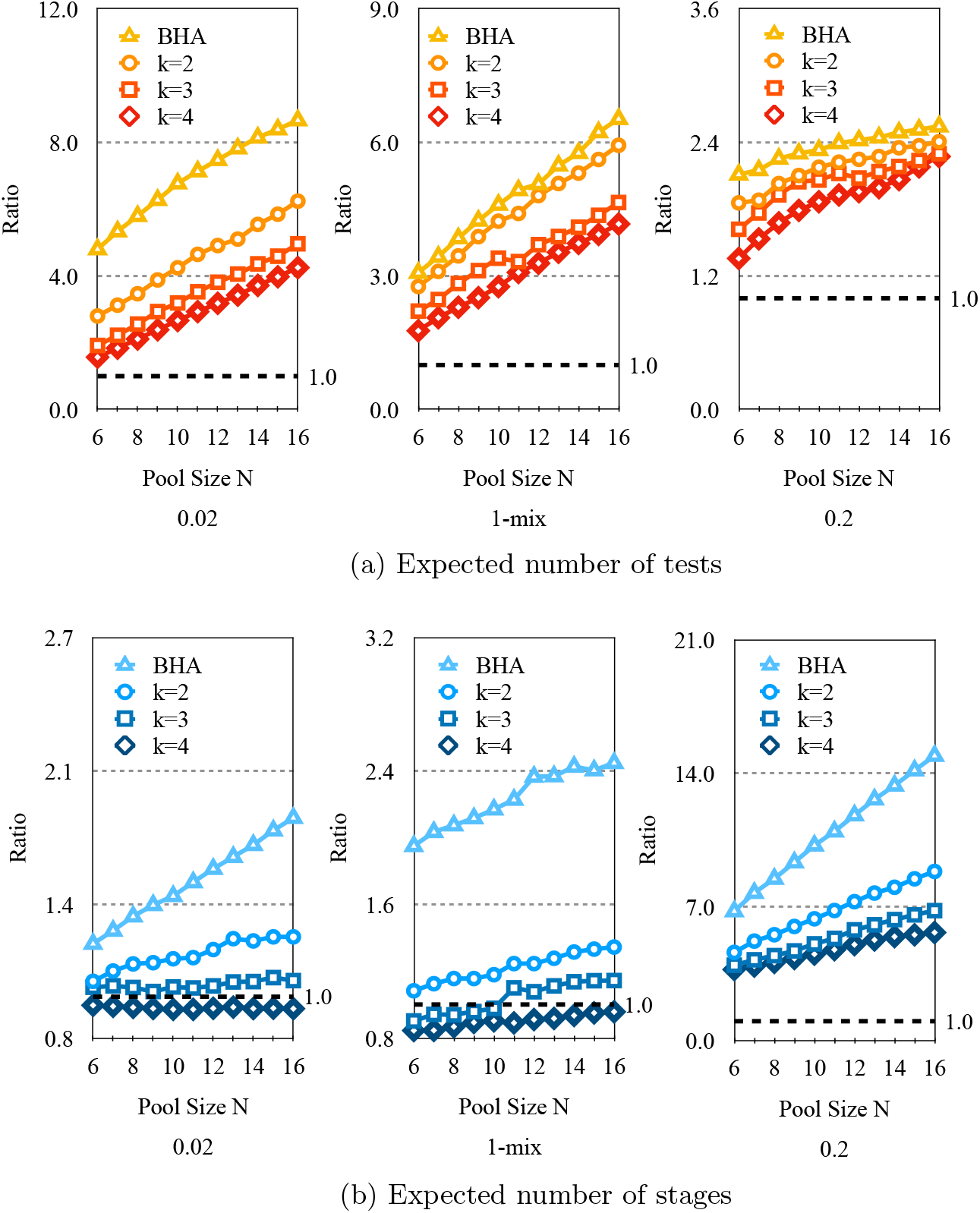
Ratio of expected number of stages, *k* = 1 to 4 versus individual testing, and expected number of tests, individual testing versus *k* = 1 to 4. Pool sizes range from N = 6 to 16 over three prior probability scenarios. Error threshold is 0.01.

Variability in the number of tests and stages per rule also is an important consideration. See Figure A.1. Note that the standard deviation in the number of stages is approximately the standard deviation for the number of tests divided by *k*, the number of tests conducted per stage. Hence, as *k* increases, the variability in number of stages can be even smaller than individual testing. Individual testing has lower variability in number of tests. The variability in number of tests and stages per algorithm appears to slightly increase in *N*. Overall, for *p*_0_ = 0.20, unless the number of stages is relative inexpensive and easy to process, individual testing becomes more attractive. We stress that a final decision depends on “local” capacity and costs.

**Example 3** In Table 2, we consider robustness with respect to the misspecification of the dilution effect and the misspecification of prior probabilities in classification. We also consider the Dorfman procedure, and the scenario when there are no dilution effects, to provide insight into the relative loss in efficiency in testing from dilution effects more generally. Except for the Dorfman procedure results, we employ the Bayesian halving algorithm (*k* = 1). Note that for *k* = 1, the number of tests and stages are the same. Three prior probability scenarios are considered (*p*_0_ = 0.02, *p*_0_ = 0.20, and 1-mix, which denotes that all prior probabilities are equal to 0.02 expect one with *p*_0_ = 0.20), as well as two different stopping thresholds (0.01 and 0.001 for negative and positive posterior classification error). In the examples, N=12. Computationally, the maximum number of stages was set to 24, and the dilution effect is the same as in Example 2. “Classification %” indicates the probability that the error thresholds have been met, through 24 stages. The first column, “Dilution”, shows performance with correct specification of the dilution effect; “No dilution” shows performance when there is no dilution effect, either in Bayesian updating or in the pooled test responses; “Misspecification of dilution effect” reflects when no dilution is assumed in Bayesian updating when actually the pooled test data does have a dilution effect; “Misspecification of Prior” reflects that the prior probabilities do not match the actual prevalence or risk of positivity, so that *p*_0_ is specified as 0.2 when the actual individual risk is 0.02, and *p*_0_ is set as 0.02 when the individual risk is 0.20; the “Dorfman” column illustrates performance with this procedure.

**Table 2:**
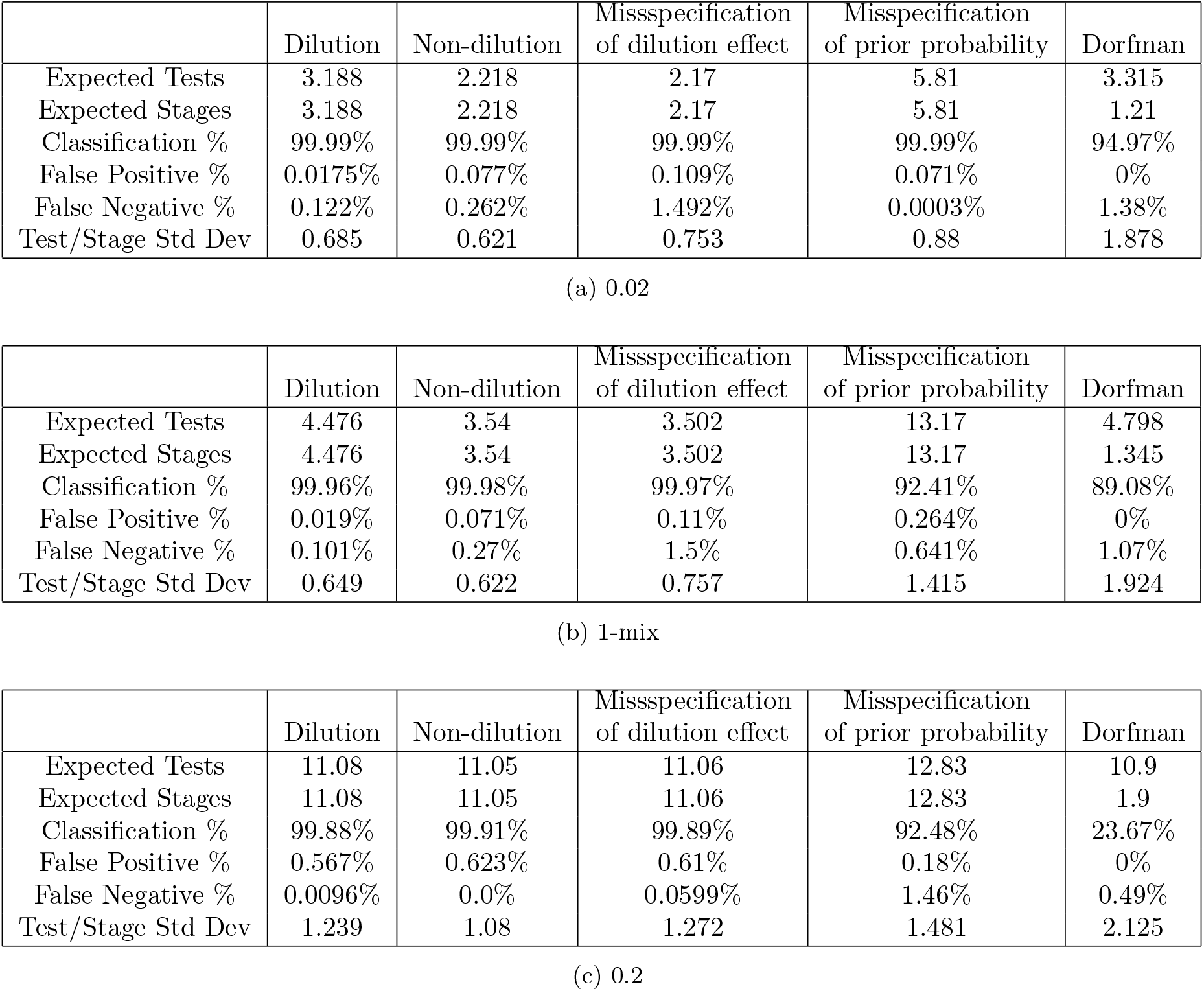
Robustness analysis of test response distribution specification and prior specification, and Dorfman procedure performance, under 3 different prior risk scenarios, with BHA.

First, we note that in comparing when the dilution effect and prior probability values are modeled correctly, versus when there is no dilution effect and probability values are modeled correctly, there can be longer test sequences under dilution effects. Interestingly, when one “ignores” the dilution effect, and assumes constant sensitivity (0.99 here), we can see higher false negative error rates, with test sequences in some cases appearing to be relatively shorter than correct modeling and having similar length to the actual non-dilution case. Another important scenario is when the prior probabilities are misspecified. Asymptotically, theoretical convergence results as described in Theorem A.2 assure that regardless of prior specification, with sufficient testing, the correct classifications can be made. However, we see here that less stringent error thresholds for stopping can lead to short sequences when risk is underspecified, and relatively higher false negative rates. These results indicate that if one is unsure about prior specification, then stringent thresholds are even more important for maintaining low error rates. We have also conducted analyses of the Dorfman procedure in the dilution effect setting, and they are presented in Table 2. Note that the Dorfman procedure either performs relatively poorly in terms of false positive and false negative rates, or does not even reach error thresholds within the maximum stage limit, so that classification rates are low. Under dilution effects, the Dorfman procedure is not generally appropriate.

Computation of performance statistics presents combinatorial challenges as *N, k* and number of stages *l* increases. To address these challenges and systematically evaluate simulations under a wider range of scenarios that are beyond the practical scope of the webtool in terms of computational time requirements, we utilize state-of-the-art big-data technologies. We have designed and developed a simulation framework based on Apache Spark Zaharia *and others* (2010). Currently, we have conducted simulations for *N* as large as 25, which involves 33,554,432 states in the lattice model. Using BHA, when *p*_0_ = 0.02 and the error threshold for stopping is 0.001, the total classification rate with a maximum number of 30 stages is 94.71512%, the false positive rate is 0.017258%, the false negative rate is 0.41134%, and the expected number of stage/tests is 5.13132. The ratios of expected number of tests and stages for this *N* = 25 scenario are approximately 8.31 (individual testing versus BHA) and 3.33 (BHA versus individual testing). For *N* = 25*p*_0_ =.02 and error threshold of 0.01, these ratios are 10.28 and 2.43. In comparing these values with those in Figure 3, where the same threshold of 0.01 was applied, note that the efficiency gains from group testing continue to increase as *N* becomes larger.

## 4. Conclusion

In a Bayesian group testing framework, dilution effects are considered that depend on pool size and the number of positives in a pool. It is seen that even when dilution effects are quite significant, asymptotically optimal strategies rely on group testing. Moreover, an intuitive Bayesian halving algorithm utilizes the natural order structure in the group testing problem that becomes explicit when viewed as a lattice model. Importantly, it is shown to attain optimal rates of convergence. *k*-test look ahead analogues of this halving algorithm also are proposed, which still can take advantage of pooling and adaptability, while reducing the number of stages in testing at the expense of testing efficiency. Computational tools have been developed that allow for assessment of the proposed group testing approaches, given inputted prior probabilities of positivity per subject, dilution effects and classification error level thresholds. These tools include the use of distributed computing to address the Big Data lattice representations for larger *N*, and they establish the feasibility and practicality of simulated group testing designs that involve performance assessments up to *N* = 25. Future work will focus on extending the computational feasibility for even larger *N*. The respective expected number of tests and stages (and standard deviations), classification error, and comparisons to individual testing are generated. Balancing the expected number of tests versus stages, as well as the respective variances and error rates, can be weighed across different *k*-test look ahead rules, and compared against individual testing. Deciding when to group test depends on the relative expense of tests versus stages, as well as the other performance statistics, and preferences may vary across testing sites.

Detailed discussion on the estimation of dilution effects is deferred here, as approaches will depend on the test response distribution model. Note that this framework can seamlessly incorporate continuous outcomes such as *C*_*t*_ values from PCR testing. In the future, it will be of interest to see if such values can provide formalized insight into the precise number of positive subjects within a pool, as opposed to just applying a binary threshold that indicates presence only. It also could be possible to consider estimating response probabilities with uncertain disease status (reflected by posterior probabilities in classification) using Bayesian latent class estimation approaches that can include nonparametric density estimation (Tatsuoka (2002); Tatsuoka *and others* (2013)). Varied and well-characterized pooled test data should become more accessible as group testing becomes more widespread through population-level surveillance. Finally, we conjecture that it will be possible to embed Bayesian halving algorithm-based testing sequences in high-volume, automated workflows, which will reduce the practical burden of requiring higher number of stages.

## Data Availability

This work does not involve data.

## 5. Software

Source code available at

https://github.com/ben072292/bayesgrouptestbackend

https://github.com/ben072292/bayesgrouptestfrontend

The web tool referenced in Section 3 is available at

https://bayesgrouptest.case.edu.

## 6. Supplementary Material

Appendix referenced in Section 1 and Section 2 is available with this paper at the Medrxiv website on Wiley Online Library. Data sharing not applicable to this article as no datasets were generated or analysed during the current study.

## Acknowledgments

This work was supported by grants NSF DRL-1561716, R01 MH65538, NSF CCF-2132049 and UL1TR002548.

