## Appendix for "BAYESIAN GROUP TESTING WITH DILUTION EFFECTS"

### BAYESIAN GROUP TESTING WITH DILUTION EFFECTS SUPPLEMENTARY MATERIALS

#### APPENDIX

##### A.1 *Notation*

We now focus on establishing the theoretical properties of Bayesian group testing with lattice models under dilution effects, and the discussion will henceforth be much more technical. We begin by reviewing the statistical formulation more formally. The set of classification states is taken to consist of the powerset of $N$ subject profiles that indicate positivity status for each of $N$ subjects. Each state in the model describes all the subjects in terms of whether each of the subjects is either *positive* or *negative* with respect to an outcome of interest. Note that there are $2^N$ possible states, corresponding to each element in the powerset. States can be identified through the subjects described as being negative, and can be partially ordered through set inclusion.

*To whom correspondence should be addressed.

© The Author 2021. Published by Oxford University Press. All rights reserved. For permissions, please

Note that state $j \geqslant k$ if the collection of negative subjects associated with state $j$ contains all the negative subjects associated with state $k$. The collection of these states thus form what will be referred to as a *powerset lattice*.

Let the collection of classification states $S$ be a finite lattice, with an unknown true state. In brief, recall that a *lattice* is a partially ordered set such that any two elements have both a unique least upper bound (*join*) and a unique greatest lower bound (*meet*). For two elements $i, j \in S$, their join and meet are respectively denoted as $i \vee j$ and $i \wedge j$. The up-set of an element $i$ is $\uparrow \mathrm{i} = \{j \in S : i \leqslant j\}$, and the down-set of $i$ is $\downarrow \mathrm{i} = \{j \in S : j \leqslant i\}$. More generally, for a set $I \subseteq S$, $\uparrow \mathrm{i} = \{j \in S :$ there exists $i \in I$ such that $i \leqslant j\}$, and $\downarrow \mathrm{I} = \{j \in S :$ there exists $i \in I$ such that $j \leqslant i\}$. A top element $\hat{1}$ is an element such that for any $i \in S, i \leqslant \hat{1}$ Similarly, a bottom element $\hat{0}$ is an element such that for any $i \in S, \hat{0} \leqslant i$. Both a top and bottom element exist in a finite lattice. For $i \in S$, define $C_i$ to be the set of covers of $i$, where $y \in S$ is a *cover* of an element $i$ if $i < y$ and there does not exist $z \in S$ such that $i < z < y$. Also, let $D_i$ denote the set of anti-covers, in other words the set of states $y \in S$ such that for $y < i$, there does not exist $z \in S$ such that $y < z < i$. In Figure 1a of the main text, note that the anti-covers for state $AB$ are the states $A$ and $B$, while state $AB$ is the only cover for both states $A$ and $B$. In general, for a powerset lattice, the covers of a true state $s$ are the states associated with one more negative subject, while its anti-covers are the states that are associated with one less negative subject. Hence, $|C_s| = N - |s|$ and $|D_s| = |s|$, where $|C_s|, |s|$, and $|D_s|$ denote respective cardinalities.

Let $\mathcal{E}$ be a finite collection of pooled tests. Denoting $X$ as the random variable being observed for an experiment $e \in \mathcal{E}$, let $f(x|e, j)$ be the class conditional probability density for $j \in S$. Note that there is a one-to-one correspondence between elements in $\mathcal{E}$ and $S \backslash \hat{0}$, so that for instance $e = j$ for some $j \in S$ implies that the set of subjects out of $N$ that are represented as negative by $j$ are the corresponding samples being pooled. Please note that here we do not use the notation $\underset{\sim}{e}$ for experiments, as in the main text. In establishing asymptotic results, assume each

$e \in \mathcal{E}$ can be replicated an unlimited number of times. Also, let $K(f, g)$ represent the Kullback-Leibler information for distinguishing distribution $f$ and $g$ when $f$ is true. Throughout, it will be assumed that for all $e \in \mathcal{E}$, there exists $j_1, j_2 \in S$ such that $K(f(\cdot|e, j_1), f(\cdot|e, j_2)) > 0$. In this case, $e$ is said to *distinguish* states $j_1$ and $j_2$. Also, assume that for any $j_1, j_2 \in S$ and $e \in \mathcal{E}$, $K(f(\cdot|e, j_1), f(\cdot|e, j_2))$ is finite.

Denote $s \in S$ as the unknown true state. For $j \in S$, let $\pi_0(j)$ denote the prior probability that $j = s$. Assume $\pi_0(j) > 0$ for all $j \in S$. Given that at the first stage a test $e_1 \in \mathcal{E}$ is selected, and a random variable $X_1$ with density $f(\cdot|e_1, s)$ has observed value $x_1$, $\pi_1(j) \propto \pi_0(j)f(x_1|e_1, j)$. Inductively, at stage $n$ for $n > 1$, conditionally on having chosen tests $e_1$, $e_2$,..., $e_{n-1}$, and having observed $X_1 = x_1, X_2 = x_2, \ldots, X_{n-1} = x_{n-1}$, a test $e_n \in \mathcal{E}$ is chosen and $X_n$ with density $f(\cdot|e_n, s)$ is observed. The posterior probability that $j = s$ then becomes

$$\pi_n(j) \propto \pi_0(j) \prod_{i=1}^{n} f(x_i|e_i, j).$$

The posterior probability distribution on $S$ at stage $n$ will be denoted by $\pi_n$.

As seen in Tatsuoka and Ferguson (2003), it is necessary and sufficient to distinguish $s$ from all other states in order for $\pi_n(s) \to 1$ almost surely. Letting $n_e$ be the number of times $e$ is administered up to stage $n$, the *limiting proportion* that $e$ is administered is denoted as $p_e$, with $p_e = \liminf n_e/n$. It is desirable to seek rules that sequentially select tests, $e_1, e_2, \ldots$, in the appropriate limiting proportions so that the posterior probability of the true state $s$, $\pi_n(s)$, converges almost surely to 1 at the fastest possible, or optimal rate. An *optimal strategy* is comprised of a collection of limiting proportions associated with each $e \in \mathcal{E}$ such that administration of the tests in their respective limiting proportions leads to convergence at the optimal rate. From Theorem A.2 in Tatsuoka and Ferguson (2003), it is argued that for $k \in S, k \neq s$,

$$\liminf_{n\to\infty} -(1/n)\log(\pi_n(k)) = \sum_{e\in\mathcal{E}} p_e K(f(\cdot|e, s), f(\cdot|e, k)).$$

The right-hand side denotes the rate at which $\pi_n(k) \to 0$ as $n \to \infty$. This rate depends on the Kullback-Leibler information values for tests that distinguish state $k$ from $s$, as well as the limiting proportions in which they are administered. Hence, larger Kullback-Leiber information values for tests are desirable, as they indicate greater discriminatory efficiency. It can be shown that $\alpha$ is the minimum of these rates among $k \neq s$, and hence maximizing $\alpha$ is equivalent to maximizing the rate of the slowest converging posterior probability terms among $k \neq s$.

##### A.2 *Dilution Effects*

Consider the following statistical formulation. Again, suppose the collection of tests $\mathcal{E}$ corresponds to all possible non-empty subsets of subjects and hence to $S \backslash \{\hat{0}\}$. In other words, every possible combination of the $N$ can be pooled and tested. For $e \in \mathcal{E}$ and $j \in S$, assume that

$$f(x|e,j) = f_{r,|e|}(x), \qquad (A1)$$

where $|e|$ denotes the number of samples in pool test $e$, and

$$r = |e| - |e \cap j|, r \geqslant 0,$$

where $r$ is the number of positive samples in pool test $e$ given $j$ is the true state.

Note when $e \leqslant j$, this implies $r = 0$. This formulation allows for response distributions to vary depending on how many positive samples are in a pool, and according to pool size. When $f_{r,|e|}$ is a Bernoulli density, let $p_{r,|e|}$ be the probability that the outcome indicates that no positive samples are present given $r$ positive samples are present and pool size is $|e|$. Note then that for $r = 0$ ( no positives), $p_{0,|e|}$ represents the specificity of a test of pool size $|e|$. Also, for $r \geqslant 1$, $q_{r,|e|} = 1 - p_{r,|e|}$ is the sensitivity of the test when $r$ positive samples are present in a pool of size $|e|$. In this section, let $K(r_1, r_2, |e|) = K(f_{r_1,|e|}, f_{r_2,|e|})$.

The following conditions, based on Kullback-Leibler information, are given to reflect the presence of dilution effects: ($i$) Suppose that both

$$K(r, r - r', |e|) \text{ and } K(r, r + r', |e|) \tag{A2}$$

are non-increasing in $|e|$ for fixed $r \geqslant 0$ and respectively for fixed $0 \leqslant r' \leqslant r$ and $r' \geqslant 0$, with $K(0, 1, |e|) > 0$ and $K(1, 0, |e|) > 0$. ($ii$) Suppose also that the Kullback-Leibler information values are non-decreasing in $r'$ for fixed $r \geqslant 0$ and $|e|$, with respectively $0 \leqslant r' \leqslant r$, and $0 \leqslant r' \leqslant |e| - r$. ($iii$) Moreover, suppose the functions in (A2) are non-increasing in $r$ for fixed $r'$ and $|e|$, with for each $r \geqslant 1$,

$$K(r, r - 1, |e|) \geqslant K(r, r + 1, |e|). \tag{A3}$$

##### A.3 *Optimal designs and optimal rates of convergence*

The following examples illustrate issues that arise in determining optimal strategies under dilution effects, which are established in Theorem A.1. An emphasis is on how the lattice structure determines the difficulty in classification. Given conditions (A1)-(A3), from a lattice-theoretic point of view, it is necessary and sufficient to distinguish a true state from its covers and anti-covers (the states that directly surround it) in order to distinguish the true state from all the others.

Example A.1 Suppose that $S$ is the lattice in Figure 1a, and that $s = \hat{0}$ (all subjects are positive), as in part (i) of Theorem A.1. The covers of $s$ are the atoms of the lattice, in other words the states that respectively represent only one of the samples being negative, $C_s = \{A, B\}$. Consider now possible strategies for distinguishing $s$ from its covers. One approach would be to individually test each subject. For instance, if sample from $A$ is tested individually, then for states in $\upharpoonleft$A$= \{A, AB\}$, since $A$ is negative, the distribution of response is $f_{0,1}$ . If the true state is in $\{\upharpoonleft \text{A}\}^c = \{\hat{0}, B\}$, then subject $A$ would be positive, and hence the corresponding distribution would be $f_{1,1}$. Hence, this test distinguishes $s = \hat{0}$ from $A$ and $AB$. Similarly, states $B$ and $AB$ can be distinguished through individually testing $B$. In sum, when testing subjects individually, the corresponding atoms are positive distinguished one test at a time. However, when response distributions depend on the number of positives in a pool, the atoms also can be distinguished from the state $s = \hat{0}$ by pooling objects. Note if subjects $A$ and $B$ are pooled, then either state $A$ or $B$ being true would result in the test having distribution $f_{1,2}$, while for $s = \hat{0}$ it would be $f_{2,2}$. This follows since if state $A$ or $B$ are true, there is one positive sample in the pool, while given $s$ is true, both sample are positive. Hence, more than one cover at a time can be distinguished from $s$ through pooled experiments. Because of dilution effects (i) and (iii), $K(1,0,1) \geqslant K(2,1,2)$, so individual testing may lead to more efficient discrimination between $s$ and its covers, while pooling $A$ and $B$ has the advantage of distinguishing $s$ from $A$ and $B$ simultaneously. Hence, in the presence of dilution effects, there is a trade-off between these testing approaches. Theorem A.1 resolves such trade-offs in terms of optimizing the rate of convergence. Note also that by dilution effect (ii), $K(2,0,2) \geqslant K(2,1,2)$, so state $AB$ is distinguished from $s$ at least as efficiently as states $A$ and $B$ when samples from $A$ and $B$ are pooled. Under individual testing, state $AB$ is distinguished whenever states $A$ or $B$ are distinguished. Hence, with either approach, $\pi_n(AB)$ will converge to zero at a rate at least as fast as either $\pi_n(A)$ or $\pi_n(B)$. □

Example A.2 In part (ii) of Theorem A.1, it is supposed that the true state $s = \hat{1}$ (all subjects are negative). In Figure 1a, state $AB = \hat{1}$. In distinguishing $s$ from its anti-covers, $D_s = \{A, B\}$, again more than one strategy must be considered. When pooling $A$ and $B$, note that the test has distribution $f_{0,2}$. States $A$ and $B$ each have class conditional distribution $f_{1,2}$. Hence, both of the anti-covers are distinguished simultaneously from $s$. It is also possible to distinguish states $A$ and $B$ by individually testing the subjects. For instance, when testing subject $A$, states $A$ and $AB$

have distribution $f_{0,1}$ since $A$ would be negative if either is the true state, while states $B$ and $\hat{0}$ would have distribution $f_{1,1}$, as $A$ is positive if either of those states is true. Hence, the anti-cover $B$ is distinguished. Similarly, the anti-cover $A$ is distinguished when $B$ is tested alone. Due to the increasing pool size, $K(0,1,1) \leqslant K(0,1,2)$, as reflected by dilution effect (i). Again, because of the presence of a dilution effect, there is a trade-off to consider in terms of distinguishing both anti-covers at once, but perhaps less efficiently than distinguishing them one at a time. As one would expect, it will be seen that as the dilution effect gets stronger, it becomes more attractive to test smaller pools. □

When $\hat{0} < s < \hat{1}$, more complex situations can arise. This is because in distinguishing covers, anti-covers can be distinguished as well when positive samples are pooled together with negative ones. However, when it is attractive to do so, there do not exist general closed form solutions of optimal strategies for such cases, as many contingencies can arise. It will instead be assumed that for

$$r \text{ positives}, 1 \leqslant r \leqslant N - |s|, \text{ and } k \text{ negatives}, 1 \leqslant k \leqslant |s|,$$

$$(r/(N-|s|))K(r, r-1, r) \geqslant (r/(N-|s|)K(r, r-1, r+k) - (k/|s|)K(r, r+1, r+k). \quad (A4)$$

As will be argued in Theorem A.1, this condition insures that for an optimal strategy, it can be assumed that the optimal value of $k$ is $k^* = 0$, and hence that asymptotically, positives and negatives are not tested together. When $r > 0$ and $k = 0$, a corresponding pooled test distinguishes covers but not anti-covers. Hence, given (A1)-(A4), when $S$ is a powerset lattice and $\hat{0} < s < \hat{1}$, it will be shown that determining optimal rates of convergence involves identifying an optimal value $r^*$, the number of positives to be pooled to distinguish covers, and $j^*$, the optimal size of pools comprised only of negatives, to distinguish the anti-covers. In contrast, when there is no dilution effect, as in Tatsuoka and Ferguson (2003), the optimal strategy is to distinguish covers by individually testing positives ($r^* = 1, k^* = 0$), and to distinguish anti-covers by pooling negatives all at once ($j^* = |s|$).

In Theorem A.1, it is assumed that the true state is known. Clearly, in practice, it is unknown, and in fact determining its identity is the primary objective of classification. Still, the following results have direct practical relevance. After, an optimal rule for selecting pooled tests will be established. In order to be optimal, a rule must first be convergent in the sense that the true state is eventually identified almost surely. In addition, it must then also eventually adopt an optimal strategy for selection of tests, corresponding to whatever state is true. Thus, Theorem A.1 characterizes the pooling sequences that must eventually be adopted almost surely under the various scenarios that can arise.

For $0 \leqslant |s| < N$, let

$$R_c^* = \sup_{1 \leqslant r \leqslant N-|s|} (r/(N-|s|))K(r, r-1, r),$$

with $r^*$ being a value that satisfies $R_c^* = (r^*/(N-|s|))K(r^*, r^*-1, r^*)$. Also, for $0 < |s| \leqslant N$, let

$$R_d^* = \sup_{1 \leqslant j \leqslant |s|} (j/|s|)K(0, 1, j),$$

with $1 \leqslant j^* \leqslant |s|$ attaining the value of $R_d^*$.

Theorem A.1 Suppose that $S$ is a powerset lattice of $N$ subjects, and let $s \in S$ denote the true state. Suppose that tests in $\mathcal{E}$ satisfy (A1), dilution effects (i)-(iii), and (A4).

(i) If $s = \hat{0}$, the optimal rate of convergence is $R_c^*$, with $|s| = 0$. This rate is attained if each possible subset of $r^*$ subjects are pooled in equal limiting proportion $1/\binom{N}{r^*}$.

(ii) If $s = \hat{1}$, the optimal rate of convergence is $R_d^*$, with $|s| = N$. This rate is attained if each possible subset of $j^*$ subjects are pooled in equal limiting proportion $1/\binom{N}{j^*}$.

(iii) Otherwise, when $\hat{0} < s < \hat{1}$, the optimal rate is

$$R_c^* \cdot R_d^*/(R_c^* + R_d^*).$$

This optimal rate is attained if each subset of $r^*$ positive subjects are pooled and tested in equal limiting proportion

$$(1/\binom{N-|s|}{r^*}) \cdot R_d^*/(R_c^* + R_d^*),$$

and all tests $e \leqslant s, |e| = j^*$ (pools of size $j^*$ consisting only of negatives) are administered in equal limiting proportion

$$(1/\binom{|s|}{j^*}) \cdot R_c^*/(R_c^* + R_d^*).$$

The respective given optimal strategies are not necessarily unique. When $j^*$ or $r^*$ are not unique, the optimal rate can be attained by any mixture of optimal allocations associated with each of the values. If $\mathcal{E}$ is restricted by bounds on pool size, Theorem A.1 can straightforwardly be extended by optimizing $j^*$ and $r^*$ with respect to the corresponding constrained values.

A.1 gives demarcations for when it is optimal to pool subjects under dilution effects. This result is in some sense analogous to that in Ungar (1960), which states that when there is no testing error with binary outcomes, it is preferable to individually test when the proportion of positive subjects is greater than $(1/2)(3-\sqrt{5})$; otherwise, group testing is preferred. In part (i) with $s = \hat{0}$ (all subjects are positive), the demarcation for when it is optimal to individually test subjects is that for all $1 < r \leqslant N - |s|$,

$$K(1,0,1) \geqslant rK(r, r-1, r). \tag{A5}$$

This condition follows by comparing rates of convergence for the covers of $s = \hat{0}$ when $r$ subjects are tested at a time. It is a regulation on dilution effect (iii). In part (ii) when $s = \hat{1}$, it is optimal to pool all the subjects if for $1 \leqslant j \leqslant N-1$,

$$K(0,1,N) \geqslant (j/N)K(0,1,j). \tag{A6}$$

More generally, it is optimal to do some form of pooling as long as $1 < j^* \leqslant N$. Condition (A6) regulates the decrease in efficiency of detecting a single positive subject versus no positive subjects as pool size increases, and hence relates to dilution effect (i). If this decrease does not occur too quickly, it is optimal to pool all subjects, which are all negative when $s = \hat{1}$. These demarcations will be illustrated below, as well as for when pooling is preferred when $\hat{0} < s < \hat{1}$.

Given the presence of dilution effects, it will be of particular interest to demarcate how strong the effects must be to alter the optimal strategies from when there is no-dilution effect. If (A5) and (A6) hold, the respective optimal strategies for when $s = \hat{0}$ and $s = \hat{1}$ correspond to the no-dilution effect case. The following examples indicate that the same optimal strategy as with no dilution effects is still optimal even in the presence of significant dilution effects.

Example A.3 Suppose $s = \hat{1}$, let $N$ be the number of objects to be classified, and suppose sensitivity decreases with pool size, but specificity stays constant. Let $f_{r,|e|}$ be Bernoulli density functions, with $p_{0,|e|} = 0.99$ for all $1 \leqslant |e| \leqslant N$, and $q_{1,1} = 0.99$. Following (A6), note that if $N = 15$, in terms of the rate of convergence, group testing all fifteen subjects is preferred if $q_{1,15} > 0.294$ and (A6) holds for the other values of $j < 15$. Further, (A6) would still be satisfied at $j = 30$ if $q_{1,30} > 0.174$. For $j = 100$, (A6) would still hold if $q_{1,100} > 0.073$. Hence, under these conditions, (A6) should hold in most practical applications.

Next, supposed that specificity is affected by pool size as well, and in equal magnitude to the sensitivity, with $q_{1,|e|} = p_{0,|e|}$. With $q_{1,1} = p_{0,1} = 0.99$ and assuming (A6) holds for all other values of $j < N$, it is still preferable to group test all subjects when $N = 15$ if $q_{1,15} = p_{0,15} > 0.689$, when $N = 30, q_{1,30} = p_{0,30} > 0.635$, and when $N = 100$ if $q_{1,100} = p_{1,100} > 0.575$.

Suppose $f_{0,|e|}$ is the density for the standard normal distribution for all $e$, and $f_{1,|e|}$ is the density for a normal distribution with mean $\mu_{1,|e|}$ and variance 1. Assume that (A6) holds for $j < N$. Letting $\mu_{1,1} = 3.0$, it can be seen that when $N = 15$, it is still more attractive to group test all objects if $\mu_{1,15} > 0.775$, when $N = 30, \mu_{1,30} > 0.548$, and when $N = 100$, if $\mu_{1,100} > 0.300$. □

Example A.4 Consider when $s = \hat{0}$. The demarcation in (A5) can similarly be illustrated numerically for when it is more attractive to individually test subjects under this case. As described in example A.2, the trade-off is between discriminatory efficiency as measured by Kullback-Leibler information versus the number of covers of $s = \hat{0}$ being distinguished per test. Pooling allows for more covers to be distinguished, but is less efficient in discrimination than individual testing. For instance, sensitivity $q_{r,r} = 0.99$ for $r \geqslant 1$, and specificity $p_{0,|e|} = 0.99$ for all $0 < |e| \leqslant N$. For this case, the demarcation is the same as in example A.3. Thus, assuming (A5) holds for all $j < N$, (A5) is also satisfied for $N = 15, 30$ and 100 when respectively $q_{14,15} < 0.294, q_{29,30} < 0.174$, and $q_{99,100} < 0.174$. This example illustrates that the dilution effect (iii) can be quite strong, and yet the no-dilution effect strategy of eventually individually testing positive subjects is not affected.

Given (A1), dilution effects (i)-(iii), and $\hat{0} < s < \hat{1}$, a simple condition that is sufficient for insuring that $r^* = 1$ is

$$K(1,0,1) \geqslant rK(r,r-1,r) \qquad (A7)$$

for $2 \leqslant r \leqslant N - |s|$. This condition is essentially the same as (A5), except (A7) applies to a smaller range of $r$. Hence, if (A5) is already established, (A7) follows as well. The numerical demarcation in example A.4 is thus applicable to (A7), and indicates that (A7) can hold generally. If the following condition holds, along with (A7), testing positives individually is optimal: for all $1 \leqslant k \leqslant |s|$,

$$(1/(N-|s|))K(1,0,1) \geqslant (1/(N-|s|))K(1,0,k+1) - (k/|s|)K(1,2,k+1). \qquad (A8)$$

Note that (A8) is a special case of (A4) with $r = 1$. Finally, given (A7) and (A8), it follows that distinguishing the anti-covers is conducted by pooled tests consisting of negatives of size $j^*$. For $1 \leqslant j \leqslant |s| - 1$, $j^* = |s|$ when

$$K(0,1,|s|) \geqslant (j/|s|)K(0,1,j). \qquad (A9)$$

Note that (A6) implies (A9). Indeed, substituting $N$ with $|s|$, note that example A.3 illustrates that $j^* = |s|$ except under strong dilution effects. In practice, checking conditions (A5) and (A6), and (A8) with $|s| = N$, is sufficient for determining for all states whether or not corresponding strategies are altered.

In sum, Examples A.3 and A.4 illustrate that dilution effects, unless severe, do not generally alter the optimal strategy of eventually pooling all negative subjects, while individually testing each of the positive subjects. One practical ramification that these results suggest is that, given low prevalence of positive samples, it is attractive to initially pool as many samples as possible, in spite of dilution effects, since the top element is most likely the true state a priori. In the next section, it will be seen that when (A5), (A6) and (A8) hold, a simple and intuitive rule for dynamically selecting pools will be optimal in the sense that optimal strategies will be selected eventually, corresponding to the unknown true state, with probability one.

##### A.4 *Optimal Pooling Selection Under Dilution Effects*

Now let us consider rules for pooling selection that, given the observed outcomes to previously administered pools and prior information, select the composition of the next stage pool to test. It is desirable to have a rule that adopts optimal strategies and attains optimal rates almost surely, no matter which state is true. It will be seen in this section that an intuitive and simple rule, a halving algorithm, attains optimal rates of convergence when the optimal strategy coincides with the no-dilution effect case, and hence even under strong dilution effects. For instance, if (A5), (A6) and (A8) hold and $s = \hat{1}$, then it is desirable for a pooling selection rule to eventually pool all objects. If $\hat{0} < s < \hat{1}$, then we would want the rule to lead to sequences of pools that eventually consist of all the negative subjects, or individually tests the positive subjects.

Theorem A.2 Under (A1), dilution effects (i)-(iii), and (A5), (A6), and (A8), for $S$ being a powerset lattice and any $s \in S$ being the true state, the Bayesian halving algorithm described by

(1) will attain the optimal rate almost surely.

##### A.5 *Bayesian Estimation of Prevalence Through Group Testing*

Based on the above-described Bayesian group testing framework, prevalence can be estimated as mixtures of posterior distributions. This approach reflects the uncertainty from test response error, and can employ pooled test results. Assume a binomial distribution. Suppose that it is of interest to estimate the proportion of positives in a target population, denoted by $\theta$, based on group testing data. Given there is uncertainty as to the positivity status of subjects, a natural estimate could rely the information provided by $\pi_n$. For instance, given a conjugate Beta prior distribution for $\theta, f(a, b)$, the posterior distribution for $\theta$ given the observed group testing outcomes is a mixture of posterior Beta densities with respect to $\pi_n$, where the Beta densities are updated depending on which state is true:

$$\sum_{j \in S} f(a + |j|, b + N - |j|) \cdot \pi_n(j).$$

For the powerset lattice, $|j|$ is the number of negatives out of the $N$ objects given state $j$ is true. Note that this mixture will converge to the posterior distribution for $\theta$ that would be obtained if the correct diagnoses for all the $N$ subjects were known exactly, given $\pi_n(s)$ converges to 1 almost surely for true state $s$.

As a simple illustration, suppose as in Example A.1 that N=2. We are interested in estimating $\theta$, and suppose a beta prior distribution, $\beta(2, 2)$. Prior mean and standard deviation are thus 0.50 and 0.22. For $n = 2$ stages, recall $\pi_2 = \{\pi_2(AB) = 0.0009, \pi_2(A) = 0.9985, \pi_2(B) = 0.0001, \pi_2(\hat{0}) = 0.0005\}$. The posterior density for $\theta$ is thus the mixture

$$(0.0009)f(4, 2) + (0.9985)f(3, 3) + (0.0043)f(3, 3) + (0.0005)f(2, 4).$$

The mixture posterior mean of $\theta$ is 0.5022, and the mixture posterior standard error is 0.1888. □

##### A.6 *Proofs*

*Proof.* of Theorem A.1

Again, let $p_e$ denote the limiting proportion that $e \in \mathcal{E}$ is administered. From Tatsuoka and Ferguson (2003), the optimal rate of convergence of $\pi_n(s)$ to 1 is thus related to the following linear program: find probability vector $\{p_e\}_{e\in\mathcal{E}}$ and $v$ to

$$\text{maximize } v$$

subject to

$$p_e \geqslant 0, e \in \mathcal{E}, \text{ and } \textstyle\sum_{e\in\mathcal{E}} p_e = 1,$$

and

$$v \leqslant \sum_{e\in\mathcal{E}} p_e K(|e| - |e \cap s|, |e| - |e \cap k|, |e|) \qquad \text{for } k \in S\backslash\{s\}. \tag{A10}$$

A probability vector, $\{p_e\}$, is sought to maximize the minimum of the right side of (A10) over $k \in S\backslash\{s\}$. Note that the right side of (A10) is the rate of convergence for state $k$ given the allocation $\{p_e\}$.

Consider first when $s = \hat{1}$. The case when $s = \hat{0}$ follows similarly. For $z \in \downarrow\!D_s\backslash D_s$, note that since $S$ is a powerset lattice, there exists $x, y \in D_s$ such that $z \leqslant x \wedge y$. Hence, $z$ is distinguished from $s$ whenever $x$ and $y$ are distinguished. Further, by (A2), the rate of convergence for terms in $\downarrow\!D_s$ is thus slowest for terms in $D_s$. We thus need only consider the terms in $D_s$ in establishing the optimal rates. Moreover, it can be established that if all experiments in $\{e : |e| = j, e \leqslant s\}$, $1 \leqslant j \leqslant N$, are each administered in limiting proportion $1/\binom{N}{j}$, the rate of convergence is $(j/N)K(0, 1, j)$. Denote such an allocation as $a(j, N)$.

It will now be shown that if

$$|e'|K(0, 1, |e'|) > |e|K(0, 1, |e|) \text{ and } |e'| < |e|,$$

then it is optimal for $p_e = 0$. Suppose $e$ is administered with limiting proportion $p_e > 0$, and let $\delta = \{p_k\}_{k\in\mathcal{E}}$ be an optimal allocation. Note that the rate of convergence contribution to each state distinguished from $s$ by administration of $e$ is $p_e K(0,1,|e|)$. However, by instead administering all $k \in \mathcal{E}$ contained in $\{\uparrow \mathrm{e}\}^c \cap D_s$, $|k| = |e'|$, in equal limiting proportion $p_e(1/\binom{|e|}{|e'|})$, the corresponding rate contribution to each state in $\{\uparrow \mathrm{e}\}^c \cap D_s$ is $p_e(|e'|/|e|)K(0,1,|e'|)$, which is greater than $p_e K(0,1,|e|)$. Hence, allocation $\delta$ can be improved, and would not be optimal.

Suppose now there exists $e$ such that $|e| < j^*$, $|e| = \inf_{k:p_k \in \delta, p_k > 0} |k|$. From above, there exists $e' \in \delta$ such that $p_{e'} > 0, |e'| > |e|, e \not< e'$, and $|e'|K(0,1,|e'|) > |e|K(0,1,|e|)$, or else the allocation is dominated. Consider

$$D' = \{\{\uparrow \mathrm{e}\}^c \ \cup \{\uparrow \mathrm{e}'\}^c\} \cap D_s = D_s \backslash \{\uparrow \mathrm{e} \cap \uparrow \mathrm{e}'\}.$$

It will now be established that the rate can be improved by administering all tests in

$$\mathcal{E}(e,e') = \{k : |k| = |e'|, \{\uparrow \mathrm{k}\}^c \cap D_s \subseteq D', \{\uparrow \mathrm{e}\}^c \cap D_s \subseteq \{\uparrow \mathrm{k}\}^c \cap D_s\},$$

in a certain equal limiting proportion given below instead of administering $e$ with proportion $p_e$. Note that there are $n' = \binom{|D'|-|e|}{|e'|-|e|}$ such experiments.

Consider two cases: First suppose

$$\textit{(i)}\ p_{e'} - p_e((K(0,1,|e|)/K(0,1,|e'|) - 1) > 0.$$

For terms in $\{\uparrow \mathrm{e}\}^c \cap D_s$, note that if tests in $\mathcal{E}(e,e')$ are administered in equal proportion

$$(p_e + p_e((K(0,1,|e|)/K(0,1,|e'|)) - 1))/n' = p_e((K(0,1,|e|)/K(0,1,|e'|))/n',$$

then the corresponding rate contribution for each distinguished state is still

$$p_e K(0,1,|e|),$$

the same as if $e$ were administered in proportion $p_e$. Moreover, administering $e'$ with proportion

$$p_{e'} - p_e((K(0,1,|e|)/K(0,1,|e'|)) - 1),$$

note that terms in $\{\uparrow \mathrm{e}'\}^c \cap \uparrow \mathrm{e} \cap D_s$ are distinguished in proportion

$$(p_e + p_e((K(0,1,|e|)/K(0,1,|e'|)) - 1))(|e'| - |e|)/(|D'| - |e|) +$$

$$p_{e'} - p_e((K(0,1,|e|)/K(0,1,|e'|)) - 1).$$

The corresponding rate contribution is greater than $p_{e'}K(0,1,|e'|)$ when $|D'| > |e'| > |e| > 0, |e'| + |e| > |D'|$, and $|e'|K(0,1,|e'|) > |e|K(0,1,|e|)$, as is assumed. Finally, note the rate contribution to states in $\{\uparrow e'\}^c \cap \{\uparrow e\}^c \cap D_s$ by administration of tests in $\mathcal{E}(e,e')$ and $e'$ is even greater.

Suppose now

$$\textit{(ii)}\ p_{e'} - p_e((K(0,1,|e|)/K(0,1,|e'|)) - 1) \leqslant 0.$$

Solving for $p_e^*$ such that

$$p_{e'} - p_e^*((K(0,1,|e|)/K(0,1,|e'|)) - 1) = 0,$$

it can be found that

$$p_e^* = p_{e'}K(0,1,|e'|)/(K(0,1,|e|) - K(0,1,|e'|)).$$

The rate contribution to terms in $\{\uparrow e\}^c \cap D_s$ from administration of experiments in $\mathcal{E}(e,e')$ in equal proportion $(p_{e'} + p_e^*)/n'$, and administration of $e$ in proportion $p_e - p_e^*$ is still

$$(p_{e'} + p_e^*)K(0,1,|e'|) + (p_e - p_e^*)K(0,1,|e|) = p_eK(0,1,|e|).$$

Further, note the rate contribution to each state in $\{\uparrow e'\}^c \cap \uparrow e \cap D_s$ by experiments in $\mathcal{E}(e,e')$is

$$(p_{e'} + p_e^*)((|e'| - |e|)/(|D'| - |e|))K(0,1,|e'|),$$

which is greater than $p_{e'}K(0,1,|e'|)$ under the given conditions. The rate contribution to states in $\{\uparrow e'\}^c \cap \{\uparrow e\}^c \cap D_s$ is thus also greater. Hence any optimal allocation will administer only tests such that $|e| = j^*$. Moreover, the optimal rate is attained when all terms in $D_s$ converge to zero at the same rate. The allocation $a(j^*, j^*)$ is thus optimal.

Suppose now that $\hat{0} < s < \hat{1}$. The condition in (A4) results from comparing the respective maximin rate among states in $C_s \cup D_s$ that would result when covers are distinguished by pools comprised of $r$ positives with no positives, versus pools with $r$ positives and $k$ negatives, and anti-covers are distinguished by $e \in \downarrow$s$, |e| = j^*$. By (A3), even when anti-covers are distinguished by pools containing both positives and negatives, it is necessary to administer tests in $\downarrow$s in positive limiting proportion. Using the arguments above, in an optimal strategy, among experiments in $\downarrow$s, only $e \in \downarrow$s with $|e| = j^*$ will be administered. Given (A4), then, among states in $C_s \cup D_s$, the maximin rate is obtained by administering $r$ positives with $k = 0$ negatives rather than if $k > 0$. Hence, for states in $C_s \cup D_s$, it is optimal to consider only strategies with $k = 0$.

If $j$ is incomparable to $s$, then there must be an object $k_1$ that is negative if $s$ is true but that is positive if $j$ is true. Let $d_1 \in D_s$ be the anti-cover associated with $k_1$ in the sense that the objects associated as negative for $d_1$ correspond to all the negatives for $s$ except $k_1$. When $d_1$ is so distinguished, so is $j$. Further, if $k = 0$ when distinguishing covers, $j$ converges at least as fast as $d_1$. Hence, only states in $C_s \cup D_s$ would determine the optimal rate, and so it is optimal to only consider strategies with $k = 0$. Following as above, only pools with $r^*$ positives and $k = 0$ negatives will be administered to distinguish covers, along with pools comprised only of $j^*$ negatives to distinguish anti-covers. Respective limiting proportions that insure that posterior probability terms for states in $C_s \cup D_s$ converge at the same rate, such as stated in (iii), are optimal.

□

*Proof.* of Theorem A.2

Define $p_j(n) = n_j/n$, where $n_j$ represents the number of times $j \in \mathcal{E}$ is administered through stage $n$, and $p_{js}(n) = n_{js}/n$, where $n_{js}$ represents the number of times $j \in S\backslash\{s\}$ is distinguished from $s$ through stage $n$. Also, let $G_{cs}$ be the minimal elements in $K_{cs}$. For a powerset lattice, $G_{cs}$ corresponds to the atom associated with the subject that is negative for $c$ but positive for $s$.

First note that the halving algorithm is convergent in the sense that $\pi_n(s) \to 1$ almost surely, and that all states are distinguished from $s$ infinitely often. Note

$$\{c\} = \uparrow c \cap \{\bigcup_{c' \in C_s \backslash \{c\}} \uparrow G_{c's}\}^c \text{ for } c \in C_s, \text{ and } \downarrow D_s = \{\uparrow s\}^c \cap \{\bigcup_{c \in C_s} \uparrow G_{cs}\}^c.$$

Also, let

$$M_n^c = |S| \cdot \pi_n(c) \text{ for } c \in C_s, \text{ and } M_n^s = |S| \cdot \sup_{j \in \downarrow D_s} \pi_n(j).$$

Define two test selection rules, procedures $A_I$ and $A_{II}$. Procedure $A_I$ is defined as follows:

$$e_n = s \text{ if } M_n^s \geqslant \pi_n(c) \text{ for all } c \in C_s\text{; otherwise, select } j \in \textstyle\bigcup_{c \in C_s} G_{cs} \text{ which maximizes } h(j, \pi_n).$$

Consider now procedure $A_{II}$:

$$e_n = s \text{ if } \pi_n(\hat{0}) > M_n^c \text{ for all } c \in C_s\text{; otherwise, select } j \in \textstyle\bigcup_{c \in C_s} G_{cs} \text{ which maximizes } h(j, \pi_n).$$

Both of these procedures attain the optimal rate of convergence for $s$ being the true state in a general lattice model. This can be seen by applying the arguments of Theorem 6 (Proposition 1) in Tatsuoka and Ferguson (2003), it should instead be stated that $a(n)/n \to B/(A+B)$). Note the slowest converging states in $S \backslash \{s\}$ are those in $C_s$ and those in $\downarrow D_s$.

When $n$ is large and hence $\pi_n(s) > 0.5$, each cover $c$ is distinguished by the corresponding element $c^* \in G_{cs}^*$. This follows since $m_n(c^*) > m_n(j)$ for any $j \in \{j' : c^* < j' \leqslant c\}$. Note also that for $n$ large, $1 - m_n(s) > 1 - m_n(j)$ for $j \in \downarrow D_s$. This implies that $h(s, \pi_n) > h(j, \pi_n)$ eventually for all $j \in \downarrow D_s$.

Again following as in Theorem 6 of Tatsuoka and Ferguson (2003), it can be argued that eventually $j \in \{\downarrow C_s\}^c$ is not administered. Further, the halving algorithm can be compared with procedures $A_I$ and $A_{II}$, establishing that the halving algorithm administers the respective optimal limiting proportions. □

##### A.7 *Addressing Computational Challenges Through Big-Data Technologies and Optimizations*

Our framework provides a self-adjustable solution for flexible parallelization that depends on workload characteristics. We have implemented several novel algorithmic optimizations to our proposed simulation framework, which achieve benefits for reducing general computational complexity of modeling and simulations, optimizing memory consumption, and providing adjustments to broad simulation scenarios based on modeling and simulation characteristics. We briefly describe a few of the major optimization techniques below:

1. Lattice models will remove subjects as soon as their individual classification threshold is reached, then reorganize remaining states and their posterior probabilities, which gradually reduces complexity associated with lattice computation and test selection rules as simulation continues.

2. We implement multiple optimizations on test selection rules by utilizing the ordered structure property of lattices in the "search" for the experiments that optimize test selection criteria. For example, for the Bayesian Halving Algorithm, if we find the posterior mass for the up-set of state $A$ is less than 0.5, then we don't need to examine states in the up-set of $A$, as they are guaranteed to have even smaller posterior mass on their respective up-sets. Such optimizations help eliminate large portions of states to be evaluated and greatly enhance the performance of proposed test selection rules.

3. We further provide two types of parallelism for test selection rules, which we refer to as "inter-lattice" and "intra-lattice". For larger lattices, we use inter-lattice parallelism where all computing resources are assigned to a single lattice to compute the test selection. For smaller lattices, we switch to intra-lattice parallelism, where each CPU core works on a single lattice. By dynamically adjusting between inter- and intra-lattice parallelism, we alleviate computation imbalance, which is an important factor in affecting performance in

an HPC setting.

4. We implement two schemes for generating simulation trees with trade-offs between speed and memory usage. The first scheme generates one big tree in parallel (single-tree scheme), which can be statistically evaluated with all true states. The second scheme generates many trees, with each corresponds to a given true state (multi-tree scheme). Here we introduce branch probabilities, in which tree branches with probabilities less than some threshold, such as 0.001 for a given state, are "pruned" from consideration in performance assessment for that state, which drastically reduces the size of trees.

5. To improve speed for the multi-tree scheme while keeping its benefit of being able to performance large-scale simulations, we propose one optimization for a frequently seen special case, which is when the individual prior probability values $p_0$ are all equal. We utilize the symmetries within the powerset lattice and Bayesian probability values, and redundant computations involving states with same number of negatives can be avoided. For instance, in such cases, when $N = 25$, computationally only 26 states need be considered, as opposed to all $2^{25}$ states.

6. The multi-tree scheme also benefits in performance improvement by skipping the evaluation of states with low prior probabilities, such as removing states with the lowest prior probability values that sum up to 1%. This has only a small effect on the accuracy of the performance statistic values.

These optimization techniques were both systematically evaluated on an Intel-based HPC with up to 476 cores and an AMD-based HPC cluster with up to 272 CPU cores.

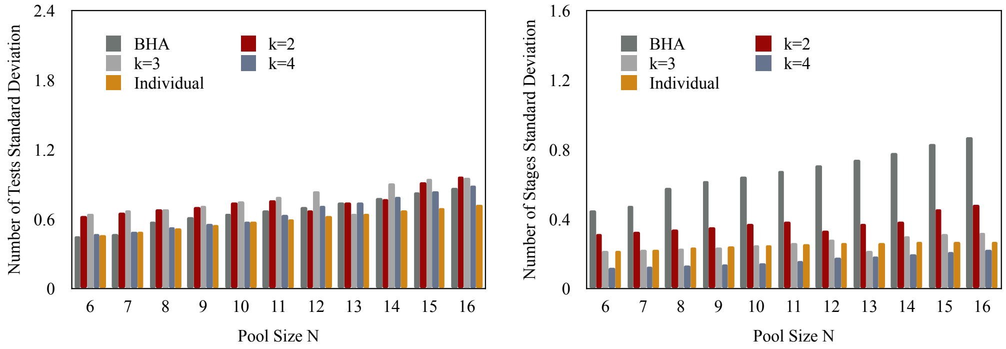


(a) 0.02

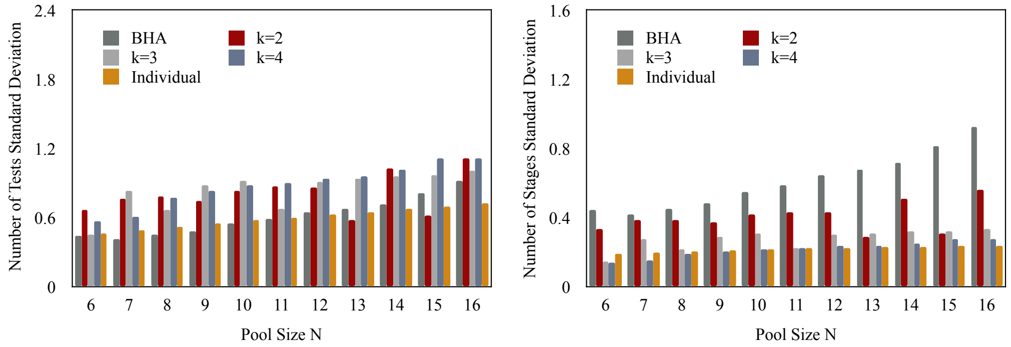


(b) 1-mix

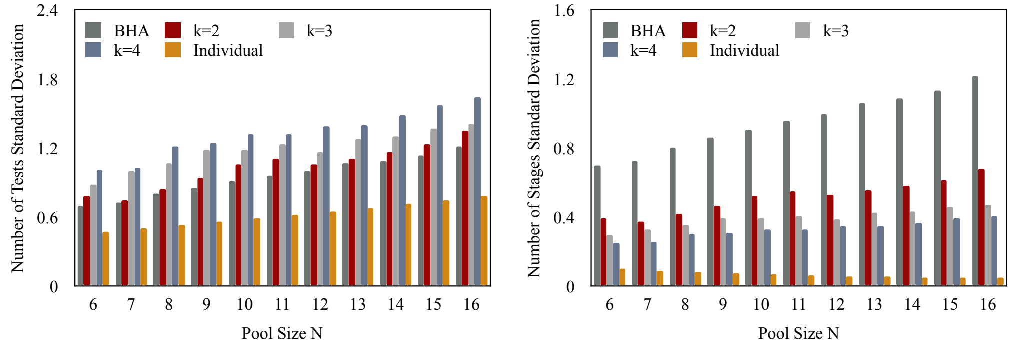


(c) 0.2

Figure A.1: Standard deviations of number of tests ranging pool size from 6 to 16 over 3 prior probability settings and k = 1 to 4.
